# The Shape of a Final Message: An Emotional Landscape in the Language of Suicide

**DOI:** 10.64898/2026.07.16.26358230

**Authors:** John P. Pestian, Daniel A. Jacobson, Ernest V. Pedapati, Eneida A. Mendonça, Benjamin H. McMahon, Julia Ive, Tracy A. Glauser

## Abstract

The emotional content of suicide notes is typically examined using categorical coding, where each labeled passage is treated in isolation from its surrounding language. In contrast, dimensional models of psychopathology propose that affective content varies along continuous gradients. We evaluated this proposition directly. Excerpts from 884 annotated suicide notes were embedded in a semantic space defined solely by their linguistic properties, and we investigated whether human-assigned emotion labels changed smoothly across this space. They did: affective tone showed clear spatial autocorrelation (Moran’s *I* = 0.18, *z* = 19.68, *p <* 0.001), an effect that replicated across three different encoders and remained after removing all within-note dependencies. Emotions occupied recognizable yet overlapping regions rather than forming distinct clusters and varied substantially in how tightly they were concentrated: love and hopelessness appeared with similar frequency, but love was far more localized (*z* = 15.7 versus 10.8). Among all emotions, hopelessness was the most linguistically diffuse, implying that a single categorical label is capturing multiple, qualitatively different manifestations of suicidal distress.

## 1 Introduction

Suicide is one of the leading causes of death worldwide, and the language people use in the final hours before they die provides a uniquely direct window into proximal suicidal thoughts [1]. Over the past fifty years, these notes have been examined using content analysis and, more recently, large-scale natural language processing [2, 3]. Throughout this work, the analytic approach has been categorical: passages are tagged with discrete emotion labels, and computational tools are trained to predict those labels.

This framework embeds an assumption that has never been tested. It treats each labeled passage as a discrete unit, counted independently of the language around it: a passage either carries the label *hopelessness* or it does not, and two passages carrying it are, for analytic purposes, equivalent. Contemporary dimensional models of psychopathology [6, 7] hold the opposite view, that affective content varies continuously, and the limits of categorical psychiatric measurement are well documented [8]. Whether the affective content of suicide notes is continuously organized or genuinely falls into discrete categories is an empirical question that categorical coding cannot answer because it presupposes the categories.

Semantic embeddings make it possible to pose this question directly. An embedding arranges linguistically similar passages close together, so that “I cannot go on like this” and “there is no point anymore” end up in neighboring locations, whereas “thank you for everything you gave me” is positioned farther away. This configuration is determined entirely by linguistic features, before any emotional labels are introduced. We can then examine whether human-provided affective annotations vary smoothly across this space or instead appear scattered without clear spatial structure. A smooth pattern would imply that affective content is continuous, as dimensional theories propose; a fragmented pattern would suggest the opposite. Since the space is defined independently of any affective tagging, this procedure evaluates the annotation scheme itself rather than simply re-encoding it.

We ask two questions of the largest fully annotated corpus of authentic suicide notes available, the Pestian *What’s in a Note* corpus [1]. First, does affective content vary smoothly across the semantic space, or does it fall into discrete clusters? Second, do individual emotions occupy separable regions, and do they differ in how compactly each is expressed?

The second question is not merely descriptive. If two emotions central to suicidal communication occupy regions of very different size, a single categorical label does not serve them equally well. Hopelessness is the case that matters most: it is among the strongest psychological predictors of eventual suicide, outperforming depression severity at ten-year follow-up [14]. A widely dispersed emotion is a candidate for containing internal sub-components that one label obscures, and dimensional accounts of suicide say what those might be. The Interpersonal Theory [4] decomposes suicidal desire into perceived burdensomeness and thwarted belonging-ness, each measured on a continuum [5]. Our corpus is not annotated for either, so we cannot test them here. But marked dispersion in a label that plausibly contains them would motivate exactly that study, which we develop as a hypothesis in the Discussion.

## 2 Results

### 2.1 The language of suicide spans a single, connected space

All 1,243 passages mapped onto one contiguous region of the embedding space, with no isolated clusters. Affective meaning shifted smoothly across it rather than appearing in discrete pockets. On a cosine *k*-nearest-neighbor graph in the original 384-dimensional space, Moran’s *I* = 0.1848 against a permuted-null mean of −0.0007 ± 0.0094 (*z* = 19.68, *p <* 0.001, 1,000 permutations): passages that are semantically near one another tend to share emotional content, and tone changes gradually with semantic position (Figure 1). The result holds across every neighborhood size tested, from *k* = 5 to *k* = 50 (*I* = 0.247 to 0.137, all *p <* 0.001; Appendix A.5).

**Figure 1:**
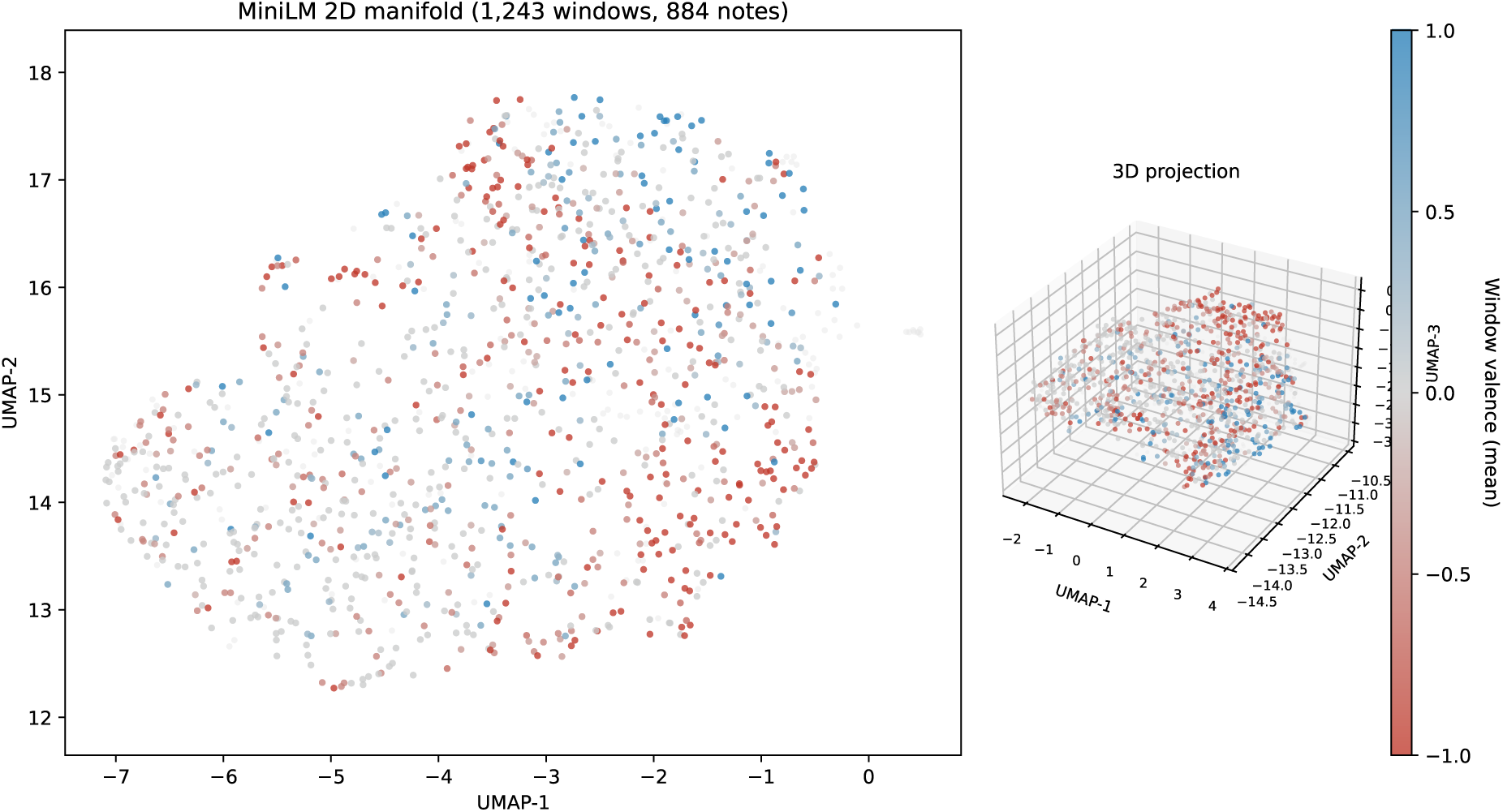
The language of suicide forms a single connected affective landscape. Two-dimensional UMAP projection of all 1,243 passage-level embeddings (primary MiniLM encoder); each point marks one passage and is colored by the affective valence of its annotated labels (positive, neutral, or negative). Affective tone is spatially autocorrelated (Moran’s *I* = 0.1848 versus a permuted-null mean of −0.0007 ± 0.0094; *z* = 19.68, *p <* 0.001; 1,000 permutations), so that tone varies smoothly with semantic position rather than separating into discrete clusters. The statistic is computed on a cosine nearest-neighbor graph in the original 384-dimensional embedding space; the projection shown here is used for display only, and was fixed prior to any affective analysis.

Four analyses establish that this is a property of the semantic space and not of any single representational choice. It is not created by the two-dimensional projection: the same statistic computed on the UMAP coordinates gives *I* = 0.1817, *z* = 16.85, *weaker* than the native-space value, so the projection loses structure rather than manufacturing it. It does not depend on the encoder: all three recover it in their own space (*I* = 0.185, 0.144, 0.139; all *p <* 0.001; Table 2). It does not depend on how structural labels are scored: excluding *information* and *instructions* from the valence definition raises the statistic to *I* = 0.1908, and testing positive-and negative-affect membership as separate binary statistics, dispensing with valence altogether, recovers the same structure (*z* = 12.37 and 9.56; Table 1). And it does not arise from the nested structure of the data: a note-blocked permutation gives *z* = 19.64, and drawing one passage at random from each of the 884 annotated notes, which removes within-note dependence entirely, yields *I* = 0.1607 ± 0.0068 across 200 draws, significant in all 200 (Appendix A.5).

**Table 1:**
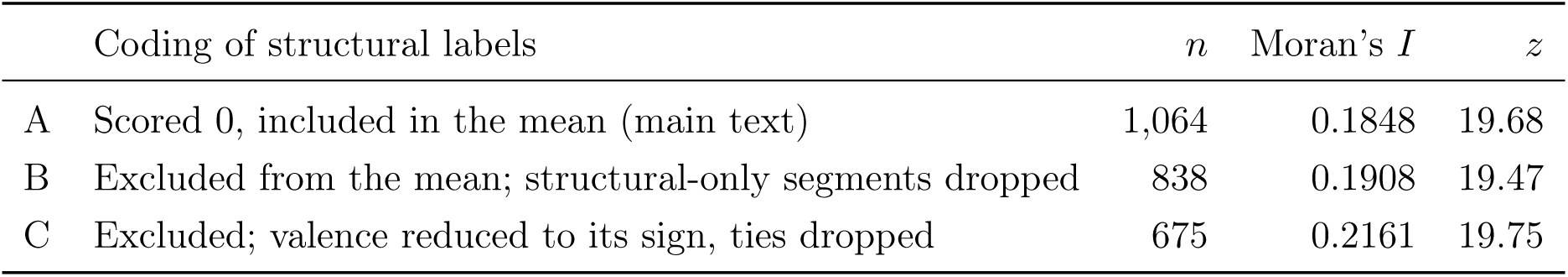
Table S3. Sensitivity of the valence spatial autocorrelation to the coding of structural labels. All rows computed in the original 384-dimensional MiniLM space on a cosine *k* = 15 nearest-neighbor graph, against a 1,000-permutation label-shuffled null. Coding A is the definition used throughout the main text.

**Table 2:** Table S1. Valence spatial autocorrelation (Moran’s. *I***) across encoders, computed in each encoder’s own embedding space.** Annotated segments (*n* = 1,064); cosine *k* = 15 nearest-neighbor graph; 1,000-permutation label-shuffled null. For reference, the same statistic computed on the 2D UMAP projection of the MiniLM embeddings gives *I* = 0.1817, *z* = 16.85; the projection slightly attenuates the effect rather than producing it.

| Encoder | Architecture (pretraining) | Dim. | Moran’s $I$ | $z$ | $p$ |
| --- | --- | --- | --- | --- | --- |
| all-MiniLM-L6-v2 | sentence-transformer (general, contrastive) | 384 | 0.1848 | 19.68 | $< 0.001$ |
| mental-roberta-base | RoBERTa-base (mental-health, MLM) | 768 | 0.1439 | 15.30 | $< 0.001$ |
| roberta-base | RoBERTa-base (general, MLM) | 768 | 0.1387 | 14.78 | $< 0.001$ |

The representation is therefore a landscape in the sense used here: a single connected domain over which affective content changes gradually, not a collection of islands.

### 2.2 Emotions occupy identifiable but overlapping regions

Twelve of the thirteen testable categories concentrated significantly: passages carrying a label appeared among their semantic nearest neighbors more often than a label-shuffled null predicts (Table 3). Ten reached *p <* 0.001; *sorrow* (*z* = 3.87) and *pride* (*z* = 3.03) were significant at a weaker threshold. Only *fear* (*n* = 35, *z* = 1.36) did not, consistent with its low base rate. Both structural categories localized strongly. The landscape is not affectively uniform: it resolves into regions that align with recognizable emotional content (Figure 2).

**Figure 2:**
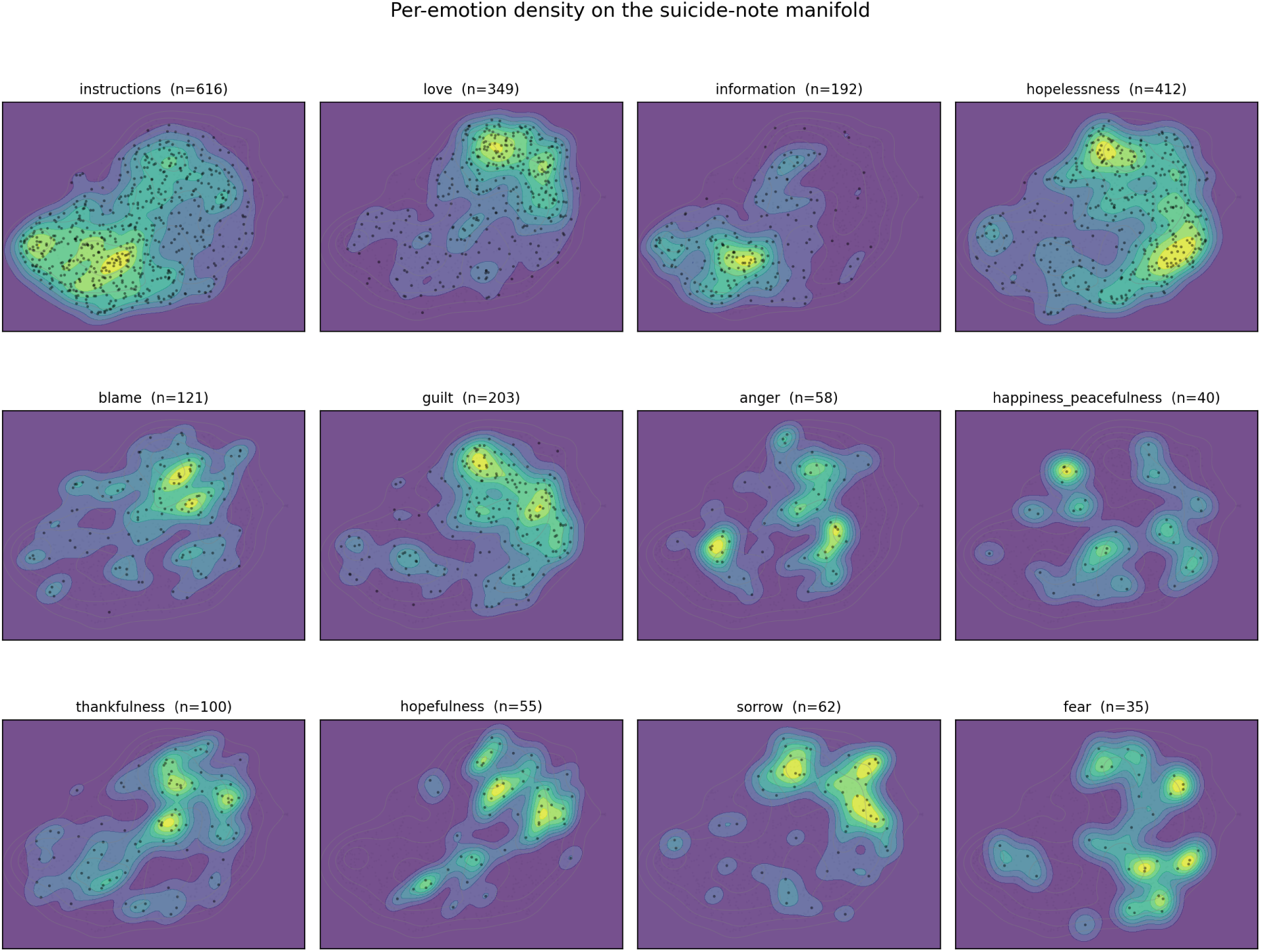
Per-emotion density across the linguistic landscape. Kernel density of labeled-passage positions for each affective and structural category with enough labeled passages to support a stable density estimate (twelve categories shown); *n* is the number of labeled passages per category, shown in each panel title. Within each panel, color indicates the local density of passages carrying that label (dark, low; yellow, high), and points mark individual labeled passages; all panels share the same underlying landscape and coordinate frame, so territories can be compared across panels. Emotions occupy identifiable but partially overlapping regions, and they differ in how compact those regions are: *love* and the other positively-toned categories concentrate in a comparatively compact, high-density territory, whereas *hopelessness*, despite being the most frequently labeled emotion (*n* = 412), spreads across a markedly larger and lower-density territory. *Pride* (*n* = 18), *forgiveness*, and *abuse* had too few labeled passages for a reliable density surface and are omitted; *pride* does concentrate significantly in the original embedding space (*z* = 3.03, *p* = 0.006) and its statistic is reported in Table 3. Panels are drawn on the UMAP projection for display; all concentration statistics are computed in the original 384-dimensional space.

**Table 3:** Table S2. Per-emotion spatial concentration (. *z***) across encoders, computed in each encoder’s own embedding space.** The rate at which a labeled segment’s *k* = 15 cosine nearest neighbors carry the same label, against a 1,000-permutation label-shuffled null. Rows ordered by descending *n*. Unmarked entries are significant at *p <* 0.001; ^∗^ denotes *p <* 0.05; “ns” denotes not significant. *Forgiveness* and *abuse* fell below the *n* ≥ 10 threshold and were not tested.

| Emotion | $n$ | MiniLM $z$ | MentalRoBERTa $z$ | RoBERTa-base $z$ |
| --- | --- | --- | --- | --- |
| instructions | 616 | 10.64 | 12.06 | 11.73 |
| hopelessness | 412 | 10.81 | 10.04 | 10.13 |
| love | 349 | 15.74 | 14.08 | 12.57 |
| guilt | 203 | 12.38 | 6.60 | 6.44 |
| information | 192 | 8.62 | 16.32 | 15.55 |
| blame | 121 | 4.28 | 5.03 | 4.36 |
| thankfulness | 100 | 6.53 | 5.19 | 3.43 |
| sorrow | 62 | 3.87* | 5.28 | 5.67 |
| anger | 58 | 6.06 | 8.66 | 8.22 |
| hopefulness | 55 | 6.07 | 4.86 | 4.81 |
| happiness_peacefulness | 40 | 6.00 | 12.28 | 12.11 |
| fear | 35 | 1.36 (ns) | 1.88* | 1.77 (ns) |
| pride | 18 | 3.03* | 2.60* | 2.24* |

The regions are identifiable but not fully separable. Of the 55 pairs of affective categories tested in the original embedding space, 33 separated at a Bonferroni-corrected threshold (*α* = 0.00091; Appendix A.5). Eight of the 22 failures involve *pride*, whose 18 labeled passages are too few to support the test. The rest are pairs applied to overlapping language. *Guilt* and *hopelessness* are the clearest case: raters assigned both labels to 21% of the passages carrying either, the highest co-labeling rate in the corpus, and even after excluding those passages, the language does not distinguish the two (*p* = 0.15). An omnibus test agrees. The silhouette coefficient of the emotion labels in the embedding space is +0.016, significantly above a shuffled-label null (*z* = 5.43) but very low in absolute terms: labels are recoverable from semantic position while explaining little of it.

Overlapping boundaries are what a continuous affective space predicts. A set of genuinely discrete categories would not produce them.

### 2.3 Love and hopelessness are captured unequally by their labels

Two categories merit closer examination. *Hopelessness* is the most heavily annotated affective category in this corpus and the leading emotion in prior content analyses of the same notes [2]; *love* is the most heavily annotated positive-affect category. Together they account for the largest share of affective annotations.

*Love* (*n* = 349) and *hopelessness* (*n* = 412) had similar local co-labeling rates (0.60 and 0.57), but concentrated very differently relative to chance: love’s effect size was substantially larger (*z* = 15.7 versus *z* = 10.8), and the ordering held in all three encoders (Table 3). Writers express love in convergent language, returning to a shared vocabulary of attachment and parting. Hopelessness, despite being the more frequently labeled of the two, is expressed across a markedly more dispersed territory (Figure 3). An emotion expressed this diffusely is a candidate for containing internal structure that a single categorical label does not capture, a point we take up in the Discussion.

**Figure 3:**
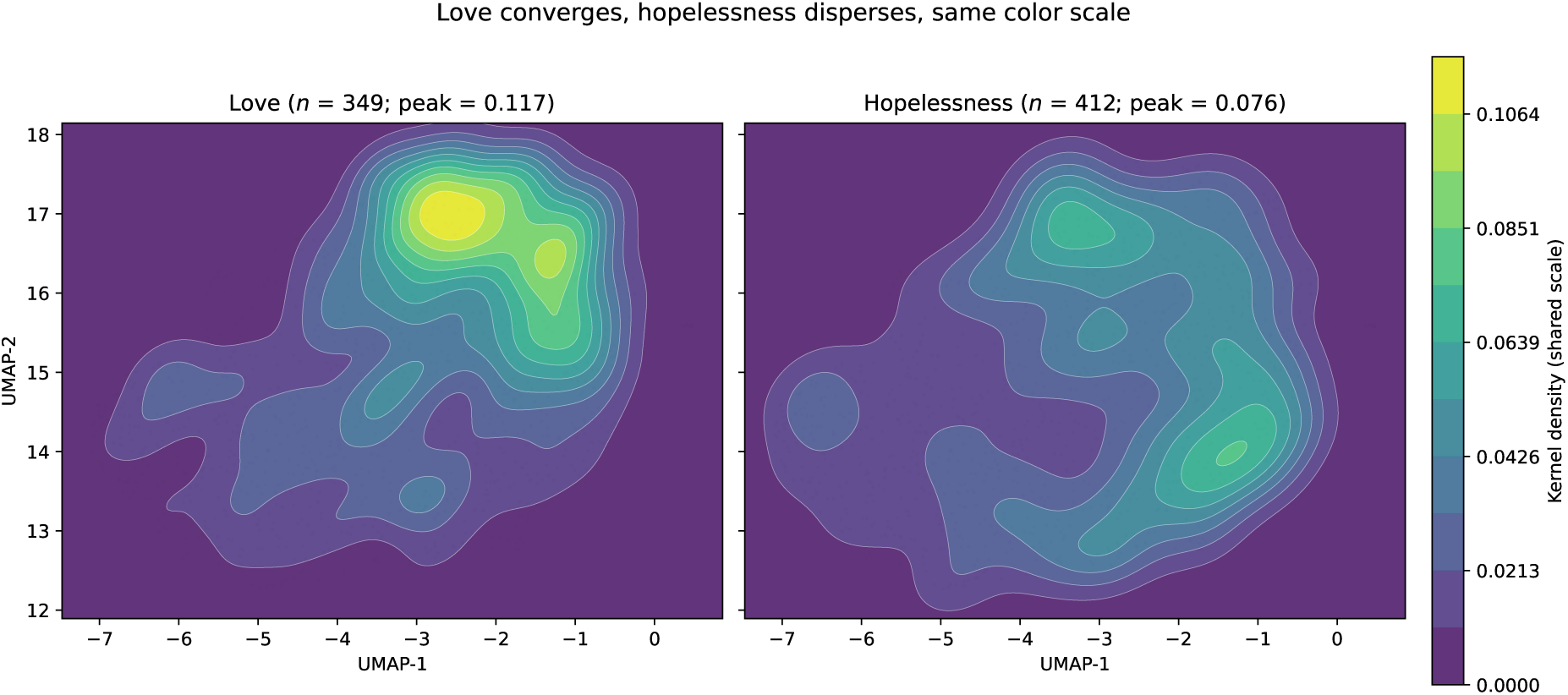
Love and hopelessness differ in linguistic concentration. Density of labeled passages across the landscape for love (left) and hopelessness (right). The two emotions have similar local co-labeling rates (0.60 and 0.57), but love’s concentration relative to a label-shuffled null is substantially stronger (*z* = 15.7 versus *z* = 10.8), indicating that love is expressed in more linguistically convergent language while hopelessness is more dispersed. Statistics are computed in the original 384-dimensional embedding space; the densities are drawn on the UMAP projection for display.

### 2.4 The findings replicate across encoders

Both headline results were recomputed under three encoders spanning two architectures and two pretraining domains, each within its own embedding space (Appendix A.6). All three recovered significant positive spatial autocorrelation at comparable magnitude, the rank ordering of per-emotion effect sizes was preserved, and the love–hopelessness asymmetry was present in each (Tables 2 and 3; Figure 4). The geometry is a property of the language of these notes rather than of any one encoder.

**Figure 4:**
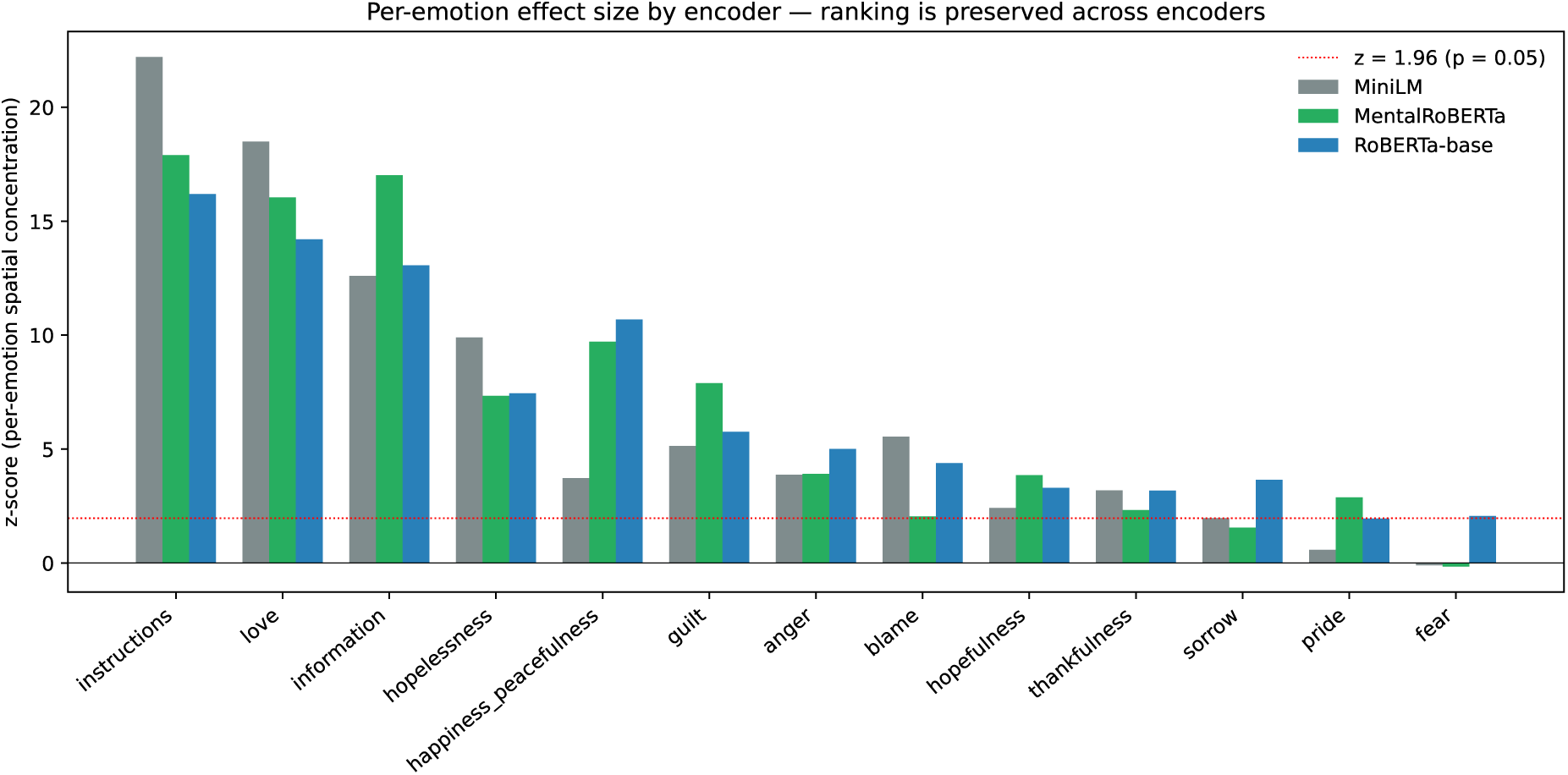
Per-emotion concentration effect size by encoder. For each emotion, the spatial-concentration *z*-score under each of three encoders (a MiniLM sentence-transformer, a mental-health-domain RoBERTa-base model, and a general-domain RoBERTa-base model), computed within each encoder’s own embedding space on a cosine *k* = 15 nearest-neighbor graph. Bars are ordered by mean *z* across encoders. The dotted line marks *z* = 1.96 (*p* = 0.05). Twelve of thirteen categories concentrate significantly under the primary encoder; only *fear* does not. The rank ordering of effect sizes is preserved across encoders, indicating that the per-emotion geometry is not specific to any one encoder family or pretraining domain. *Love*concentrates most strongly of the affective categories under all three encoders, and more strongly than *hopelessness* in each, despite *hopelessness* carrying more labels.

## 3 Discussion

Two findings emerge. Affective content varies smoothly across a semantic space whose structure is fixed by language alone, rather than falling into discrete clusters. And emotions differ in how consistently they are expressed: love is linguistically convergent, whereas hopelessness, labeled at least as often, is distributed across a substantially broader and more diffuse territory.

### 3.1 Affective content in suicide notes varies smoothly

Affective tone showed clear spatial autocorrelation in the semantic space itself (Moran’s *I* = 0.1848, *z* = 19.68). The pattern held across three encoders spanning two architectures and two pretraining regimes, across every neighborhood size tested, under three alternative codings of structural language, and after the removal of all within-note dependence. It is therefore not a property of one language model, not an artifact of the two-dimensional projection used for the figures, and not a consequence of treating multiple passages from one note as independent.

This is a direct empirical test, in the writers’ own language, of the assumption underlying dimensional models of psychopathology [6, 7], and one that categorical coding cannot perform, because it presupposes what it would need to test. Affective content does not fall into separate discrete categories; it varies continuously across neighboring regions of meaning. Categorical labels are not wrong. The categories they name are genuinely present, and twelve of thirteen concentrate significantly. But they are incomplete, because they discard the connected structure that organizes and links the regions they label, and because their boundaries overlap: only 33 of 55 category pairs are separable, and the emotion-label silhouette, though significant, is close to zero.

### 3.2 Emotions differ in how well single labels capture them

Love and hopelessness are labeled at similar rates among semantic neighbors, yet love concentrates far more strongly relative to chance. Writers express love convergently, returning to a shared vocabulary of attachment and parting, so the label fits its territory well. Hopelessness is expressed diffusely, across a much larger region. The label *hopelessness* therefore functions differently from *love*: it designates a broad, internally heterogeneous territory rather than a compact one, and any analysis treating one hopelessness-labeled passage as exchangeable with another averages over considerably more linguistic variability than the same operation performed on love.

The instruments used to assess hopelessness and proximal suicide risk, including the Beck Hopelessness Scale [13], item nine of the PHQ-9 [15], and the Columbia Suicide Severity Rating Scale [16], treat hopelessness as a unitary construct scored on a single severity dimension. The diffuse territory the label occupies here raises the possibility that two patients endorsing hopelessness at the same level are communicating different states. We do not claim this study demonstrates that. Establishing it would require linking semantic position to independent clinical assessment, which we did not do.

### 3.3 A hypothesis: structure within hopelessness

Why is hopelessness so dispersed? One possibility is that it is simply a broader lexical category. A more specific hypothesis follows from dimensional theories of suicide. The Interpersonal Theory [4] holds that what is clinically called hopelessness is not one state, distinguishing perceived burdensomeness, the conviction that one’s death would benefit others, from thwarted belongingness, the experience of social disconnection, each measured on a continuum [5]. If passages expressing these constructs are all coded *hopelessness* under a schema that does not contain them, dispersion is precisely what one would expect.

An exploratory analysis (Appendix A.9) sharpens the hypothesis and constrains it. Hopelessness does not partition into discrete sub-types: density-based clustering of the 412 hopelessness-labeled passages returns no clusters at all, and the best *k*-means silhouette across *k* = 2 to 6 is 0.09, low enough to indicate that any partition is imposed rather than found. Within that continuous territory, however, variation is lexically structured. One pole is hopelessness expressed in relational terms, marked by address, endearment, apology, and forgiveness, lying very close to the love territory and co-labeled *love* in half its passages. The other is somatic and durational, marked by illness, deterioration, and elapsed time, and lying markedly further from love. The relational pole carries the surface features that the Interpersonal Theory associates with perceived burdensomeness.

We frame this as a hypothesis, and the clustering result tells us what kind of hypothesis it can be. The corpus is annotated with a schema containing neither burdensomeness nor belongingness, so nothing here identifies those constructs; and the absence of discrete clusters argues positively against a sub-type account. What the geometry supports is the weaker claim that hopelessness varies continuously along an axis a dimensional theory predicts, which is what such a theory should want. Testing it requires a dedicated study: a secondary annotation pass coding hopelessness-labeled passages for the relevant constructs, then a test of whether they occupy distinguishable positions along the observed gradient.

### 3.4 What follows, and what does not

The dispersion finding has one methodological implication. Detection systems trained on prototypical expressions of an emotion perform well when that emotion is expressed convergently and less well when it is not. Hopelessness, which is central to suicidal communication, falls into the latter case; prior work shows that when a classifier cannot sharply separate cases from controls, standard calibration loses resolution and more flexible strategies are needed [12]. Representations that locate a passage within a continuous semantic space, rather than matching it to a template, are better suited to emotions whose expression does not converge.

Beyond that, we make no clinical claim. Nothing here demonstrates that semantic position predicts outcome, that the sub-regions described above are separable in living patients, or that any change to assessment practice would improve risk stratification. These notes were written by people who died, and the prudent direction of inference runs from their language toward better-designed prospective work, not toward the clinic. The study this geometry motivates is a prospective one, linking semantic position in a patient’s own words to theory-guided constructs and to outcome. Until that study is done, what this paper offers the clinician is a reason to attend to *which* kind of hopelessness a patient expresses, and not only how severely.

### 3.5 Limitations

Several limitations qualify these conclusions.

The most consequential concerns what the geometry measures. The embedding is derived from words, and the annotations were assigned by humans reading those same words. Passages containing similar vocabulary will therefore embed near one another *and* attract similar labels, and the observed spatial autocorrelation may partly reflect annotators responding to lexical surface features rather than the continuous organization of affect itself. The two are not fully separable in an observational design of this kind. Relatedly, we report Moran’s *I* against a random-permutation null, which establishes that the observed arrangement is not chance, but we have no reference model for the value a genuinely discrete category system would produce under the same pipeline. The magnitude of *I* = 0.18 should therefore be read as significantly non-random rather than as calibrated against a discreteness benchmark.

The corpus comprises notes from people who died by suicide *and* left a note; note-leavers differ systematically from the majority who do not [11], so these findings should not be generalized without further work. The material is historical and primarily in US English, and the linguistic structure of contemporary suicidal communication may differ. The analysis rests on an existing 15-emotion schema: the reported patterns describe how those labels are arranged, and do not validate the labels themselves. The design is cross-sectional at the level of authorship, so we cannot say whether the patterns reflect stable individual profiles or transient crisis states. Finally, *fear* did not reach significance and *pride* and *sorrow* were significant only at a weaker threshold; given their low base rates, their spatial configuration should be regarded as indeterminate rather than absent.

### 3.6 Conclusion

The emotional content of suicide notes forms a continuous, smoothly structured linguistic space rather than a set of discrete categorical clusters. Emotions occupy identifiable but overlapping territory within it, and they differ markedly in how tightly they concentrate. The broad dispersion of hopelessness, against the convergence of love, indicates that one categorical label can span a far wider and more heterogeneous linguistic territory than another, and motivates, at a hypothesis-generating level, the possibility that hopelessness absorbs several distinct forms of suicidal distress. What the writers’ own words most directly support is the structure of the space itself: the language of suicide does not divide into bins, and representations that respect this can reach a resolution that categorical coding cannot.

## 4 Methods

### 4.1 Corpus and ethics

The Pestian *What’s in a Note* corpus comprises 1,278 notes authored by individuals who died by suicide, collected over the period 1950–2012 by Edwin Shneidman of UCLA, Antoon A. Leenaars, and John Pestian of Cincinnati Children’s Hospital Medical Center [1]. The present study analyzes the 884 notes for which complete consensus annotations were available. Each note was independently labeled by at least three of approximately 160 trained volunteer annotators, recruited from communities of individuals bereaved by suicide (“survivors of suicide loss”), 18 of whom were mental health professionals and the remainder of whom held educational qualifications ranging from a high-school degree to a professional doctorate [2]. All annotators completed a structured training protocol, including practice annotation of 10 reference notes against a gold standard, before contributing to the corpus. Agreement between the volunteer annotators and clinician annotation of the same material was 78% [1]. Notes with initial inter-annotator disagreement were resolved through a structured arbitration phase to produce the consensus labels used in this analysis [2]. The 15-category coding framework includes thirteen affective categories (*love, thankfulness, hopefulness, happiness_peacefulness, pride, forgiveness, hopelessness, guilt, blame, anger, sorrow, fear, abuse*) and two structural categories (*information, instructions*) [2]. Notes were collected and de-identified under IRB-approved procedures at Cincinnati Children’s Hospital Medical Center (IRB 2016-1505). The surviving family members of decedents provided consent for these materials to be used in research.

### 4.2 Constructing the linguistic landscape

The analytic approach has three conceptual steps; the technical parameters of each are given in Appendix A so that the main text remains readable without them.

*From notes to passages.* Individual notes are not affectively uniform, and the corpus annotations reflect this: raters assigned emotion labels at the span level, and among notes long enough to include at least two annotated spans, the majority (71%) contain more than one distinct affective category. If we treated each note as a single unit, we would be averaging away exactly this within-note variability. We therefore segmented each note into passages of at most 800 characters, with 200 characters of overlap between consecutive passages. Passages are long enough to support a stable semantic representation but short enough to capture a single affective episode instead of merging several; the modest overlap ensures that affective content near a passage boundary is not split between two passages. This produced 1,243 passages from 884 notes (median 1 per note, maximum 16), of which 1,064 had at least one rater annotation. Because 706 notes were shorter than 800 characters, they yielded a single passage each, and the mean passage length across the corpus was 471 characters. A passage can receive multiple labels, so per-emotion frequencies sum to more than the total number of annotated passages.

*From passages to positions.* Each passage was transformed using a pretrained sentence-transformer [9] into a numerical encoding of its semantic content, such that passages with similar wording receive similar encodings. These encodings define the *landscape*: passages expressing similar ideas lie close to one another in it. All statistical tests reported in this paper are computed in this original encoding space. For display, the encodings are additionally projected to two dimensions [10]; the projection is used for figures only, and no result depends on it. Both the encoding and the projection were fixed prior to any affective analysis, so the tests evaluate whether affective content is structured over a space whose configuration was determined purely by linguistic information.

*Affective tone of a passage.* For the affective tone analysis, each annotated passage was assigned an overall valence. Positive-affect labels (*love, thankfulness, hopefulness, happiness_peacefulness, pride, forgiveness*) scored +1; negative-affect labels (*hopelessness, guilt, blame, anger, sorrow, fear, abuse*) scored −1; structural labels scored 0. A passage’s valence is the mean over its labels. Because the treatment of structural labels is a substantive choice, three alternative codings are reported in Appendix A.5.

### 4.3 Testing for smooth affective variation

The hypothesis that the language of suicide forms a connected space with smooth affective variation, rather than discrete categorical clusters, yields a testable implication: passages close to one another in the semantic space should, on average, share affective tone. We evaluated this with Moran’s *I*, a standard measure of spatial autocorrelation, computed on a cosine *k*-nearest-neighbor graph in the original embedding space and compared against a null in which valence is permuted across passages while the graph is held fixed (Appendix A.5). A positive, significant Moran’s *I* implies that affective tone changes gradually with semantic position; a non-significant result would imply that it does not.

### 4.4 Locating individual emotions

For each affective category with at least 10 labeled passages, we tested whether passages carrying that label appeared among their semantic nearest neighbors more often than a label-shuffled null predicts (Appendix A.5). To assess whether emotions occupy *separable* territory rather than merely each being individually concentrated, we tested every pair of affective categories for nearest-neighbor label purity in the embedding space against a label-shuffled null, with Bonferroni correction across the 55 pairs. As an omnibus check on the label structure as a whole, we computed the silhouette coefficient of the emotion labels among single-label passages, against a shuffled-label null. *Forgiveness* and *abuse* had too few labeled passages to test and were omitted.

### 4.5 Robustness and reproducibility

Every headline result was recomputed under three encoders spanning two model architectures and two pretraining domains: a MiniLM sentence-transformer (primary), a mental-health-domain RoBERTa-base model, and a general-domain RoBERTa-base model, in each case within that encoder’s own embedding space (Appendix A.2, A.6). Three further robustness analyses are reported in the Appendix: sensitivity to the neighborhood size *k*, to the coding of structural labels, and to the nested structure of the data (Appendix A.5); stability of the display projection across 20 random initializations (Appendix A.7); and sensitivity to the de-identification substitution tokens present throughout the corpus (Appendix A.8).

### 4.6 Use of AI tools

During preparation of this manuscript, the authors used Claude (Anthropic) to assist with manuscript editing and restructuring and with drafting analysis code. The authors reviewed and verified all content, analyses, and conclusions, and take full responsibility for the work. No generative AI was used to produce any figures or images.

## Data Availability

The corpus contains the final written communications of individuals who died by suicide and is therefore subject to substantial protections beyond standard de-identification. Access requires (i) an approved research protocol from the requesting investigator's institutional review board, (ii) a data use agreement with Cincinnati Children's Hospital Medical Center governing permitted uses, storage, sharing, and disposition of the data, and (iii) documentation of the requesting investigator's institutional affiliation and prior research experience in suicidology, clinical psychology, or a closely related field. These conditions reflect the corpus's status as the final words of decedents and the corresponding obligations to their surviving family members, who consented to research use on the understanding that the materials would be handled with appropriate care. Inquiries should be directed to the corresponding author. Because the analyses operate directly on this restricted corpus, full computational reproduction requires access to the corpus under the conditions above; no derived data product that would permit reconstruction of the underlying note text is released.

## Acknowledgments

We thank the families who consented to the use of their loved ones’ final words for research. The notes analyzed here were written by people in the most desperate moments of their lives, addressed in most cases to those who loved them; this research is possible only because grieving family members chose to share what was meant for them alone. This work is a debt to them, and we have tried to honor it by treating the words themselves, and the clinical truths those words contain, as the primary contribution. We also acknowledge the foundational role of the late Edwin Shneidman of UCLA, whose decades-long collection of suicide notes constitutes a substantial portion of the historical material on which this corpus rests, and of Antoon A. Leenaars, whose body of work on the linguistic and psychological structure of suicide notes shaped the conceptual ground for studies of this kind.

The manuscript was coauthored by UT-Battelle, LLC under Contract No. DE-AC05-00OR22725 with the US Department of Energy. The US Government retains and the publisher, by accepting the article for publication, acknowledges that the US Government retains a nonexclusive, paid-up, irrevocable, worldwide license to publish or reproduce the published form of this manuscript, or allow others to do so, for US Government purposes.

## Funding

This work was funded by Cincinnati Children’s Hospital Medical Center as part of our Pursuing our Potential Together initiative.

## Author contributions

Author contributions are reported using the CRediT taxonomy. J.P.P.: Conceptualization; Data curation; Formal analysis; Funding acquisition; Methodology; Project administration; Software; Supervision; Visualization; Writing – original draft; Writing – review & editing. D.A.J.: Methodology; Validation (verification of the formal analysis); Writing – review & editing. E.V.P.: Validation (clinical interpretation of the affective categories and suicide-note material); Writing – review & editing. E.A.M.: Validation (clinical interpretation); Writing – review & editing. B.H.M.: Conceptualization; Validation (verification of the formal analysis). J.I.: Methodology; Writing – review & editing. T.G.: Methodology; Validation; Visualization; Writing – original draft; Writing – review & editing. All authors reviewed and approved the final manuscript.

## Conflicts of interest

The authors declare no competing interests.

## Ethics statement

All notes were collected and de-identified under IRB protocols at Cincinnati Children’s Hospital Medical Center (IRB 2016-1505). Analyses used only de-identified text. No information capable of identifying any decedent or any surviving family member is presented anywhere in this manuscript or its supplementary materials. The authors approached this work with the awareness that each note represents an individual life and an individual loss, and that no analytic choice should compromise the dignity of the writers or the privacy of their families. No verbatim passage from any note is reproduced in this manuscript.

## Data availability

The Pestian *What’s in a Note* corpus contains the final written communications of individuals who died by suicide and is therefore subject to substantial protections beyond standard de-identification. Access requires (i) an approved research protocol from the requesting investigator’s institutional review board, (ii) a data use agreement with Cincinnati Children’s Hospital Medical Center governing permitted uses, storage, sharing, and disposition of the data, and (iii) documentation of the requesting investigator’s institutional affiliation and prior research experience in suicidology, clinical psychology, or a closely related field. These conditions reflect the corpus’s status as the final words of decedents and the corresponding obligations to their surviving family members, who consented to research use on the understanding that the materials would be handled with appropriate care. Inquiries should be directed to the corresponding author. Because the analyses operate directly on this restricted corpus, full computational reproduction requires access to the corpus under the conditions above; no derived data product that would permit reconstruction of the underlying note text is released.

## Code availability

The analysis code is available from the corresponding author on reasonable request. The full computational methodology, including the named embedding models, the valence assignment of emotion categories, the projection parameters, and the statistical tests, is described in the Methods and Appendix A. Because executing this code requires the IRB- and DUA-restricted corpus described under Data availability, the code is provided for inspection rather than independent re-execution.

## **A** Technical Appendix: Methods Detail

This appendix gives the technical specifications of the analysis, in the order the analysis was carried out: from the corpus to passages, to their semantic encoding, to the projection used for display, to the statistical tests, to the robustness analyses, and finally to the software environment. The Methods section explains the same procedures conceptually; nothing here alters how any result should be interpreted.

### **A.1** Passage segmentation

Each note was divided into segments of at most 800 characters, with a 200-character overlap between consecutive segments (equivalently, a 600-character step). This yielded 1,243 segments from 884 notes (median 1 segment per note, maximum 16); 1,064 segments contained at least one rater annotation. Because 706 of the 884 notes were shorter than 800 characters, each produced a single truncated segment; the mean segment length across the corpus was 471 characters, and segments at the full 800-character width correspond to roughly 130–160 English words. Adjacent segments within a note share 25% of their text by construction. No analysis in this paper depends on the ordering or adjacency of segments; every test treats segments as an unordered set, so this overlap does not enter any reported result.

### **A.2** Semantic encoding

Each passage was transformed into a fixed-length vector using one of three pretrained transformer encoders, chosen to span two architectures and two pretraining domains (Section A.6). No encoder was fine-tuned on the corpus; passages were passed through each in inference mode only.

The *primary encoder* was all-MiniLM-L6-v2 [9], a 6-layer sentence-transformer trained on more than one billion sentence pairs from paraphrase, question-answering, and natural-language-inference corpora, optimized by contrastive learning to produce 384-dimensional embeddings in which semantically similar passages lie close together. This encoder is the substrate for all primary analyses.

The *mental-health-domain encoder* was mental-roberta-base [19], a RoBERTa-base model continually pretrained on mental-health text from Reddit communities concerned with depression, anxiety, and suicidal ideation. It produces 768-dimensional embeddings; passage vectors were obtained by mean-pooling over token embeddings.

The *general-domain encoder* was roberta-base [20], trained on BookCorpus, English Wikipedia, CC-News, OpenWebText, and Stories. It produces 768-dimensional embeddings, mean-pooled as above.

Together these allow the findings to be evaluated under a domain-matched encoder, an architecturally equivalent general-domain encoder, and a sentence-transformer trained for semantic similarity.

### **A.3** Projection

All statistics in this paper are computed in the encoders’ original embedding spaces (384 dimensions for MiniLM, 768 for the two RoBERTa variants), not in any projection. The projection serves one purpose: display. Encoded vectors were projected to two dimensions using UMAP [10] with *n*_neighbors = 25, min_dist = 0.15, cosine metric, and a fixed random seed (42). This two-dimensional embedding is the substrate for every figure and for no reported statistic. It was generated once, prior to any affective analysis, and then held fixed.

For completeness, we computed trustworthiness and continuity [17] for the primary encoder’s projection at *k* = 15: trustworthiness 0.83, continuity 0.88. These figures characterize the fidelity of the display and do not bear on any result, since no result is computed on the projected coordinates.

### **A.4** Per-segment valence

Each annotated segment was assigned a valence equal to the mean of its label scores: positive-affect labels (*love, thankfulness, hopefulness, happiness_peacefulness, pride, forgiveness*) scored +1, negative-affect labels (*hopelessness, guilt, blame, anger, sorrow, fear, abuse*) scored −1, and structural labels (*information, instructions*) scored 0. Sensitivity to the last of these choices is reported in Section A.5.

### **A.5** Spatial autocorrelation and concentration statistics

All statistics in this section are computed on a *k*-nearest-neighbor graph built in the encoder’s original embedding space using cosine distance. No statistic is computed on UMAP coordinates. Unless stated otherwise, *k* = 15 and the encoder is MiniLM.

*Landscape spatial autocorrelation.* Moran’s *I* was computed on per-segment valence over the 1,064 annotated segments. The null was obtained by permuting valence across segments 1,000 times with the neighbor graph held fixed. The observed value was *I* = 0.1848 against a null of −0.0007 ± 0.0094 (*z* = 19.68; no permutation reached the observed value, *p <* 0.001).

*Neighborhood-size sensitivity.* Recomputing at *k* ∈ {5, 10, 15, 25, 50} gives *I* = 0.2467, 0.1990, 0.1848, 0.1630, 0.1369 (*z* = 15.84 to 28.00; all *p <* 0.001). The statistic is monotone in *k*, as expected for widening neighborhoods, and significant throughout.

*Comparison with the projected space.* The identical statistic computed on the 2D UMAP coordinates gives *I* = 0.1817, *z* = 16.85. The projected value is lower than the native value at every *k* tested, indicating that the projection loses a small amount of the structure present in the embedding space rather than creating it.

*Nested data structure.* Segments are nested within notes, and 178 notes contribute more than one. Two analyses address the resulting non-independence. A note-blocked permutation, in which whole-note runs of valence are shuffled between notes so that within-note structure is preserved under the null, gives *z* = 19.64 against the standard null’s *z* = 19.68. A one-window-per-note bootstrap, drawing a single segment at random from each of the 804 notes that contribute a valence-labeled segment and recomputing the statistic, gives *I* = 0.1607 ± 0.0068 (range [0.1445, 0.1842]) across 200 draws, all 200 significant at *p <* 0.05 against a 500-permutation null. Neither materially changes the result.

*Per-emotion concentration.* For each category with *n* ≥ 10 labeled segments, a concentration statistic was computed as the rate at which a labeled segment’s *k* = 15 nearest neighbors in the embedding space also carry that label, against a null of 1,000 label-shuffled draws. Results for all three encoders are in Table 3. Kernel density estimates of segment position, used only for the density figures, employ a Gaussian kernel with Scott’s-rule bandwidth on the 2D projection. *Pairwise emotion separability.* Density-overlap methods do not transfer to a 384-dimensional space. Separability was therefore tested by nearest-neighbor label purity. For each pair of affective categories (*A, B*) we took the segments labeled *A* but not *B* and those labeled *B* but not *A*, built a *k* = 15 cosine neighbor graph among that subset in the embedding space, and computed the fraction of neighbor relations joining two segments of the same label, against 1,000 permutations of the *A/B* label within the subset. Of the 55 pairs, 33 were separable at a Bonferroni-corrected threshold of *α* = 0.05*/*55 = 0.00091. Eight of the 22 non-separable pairs involve *pride* (*n* = 18). The strongest substantive non-separability is *guilt* versus *hopelessness* (purity 0.648, *z* = 1.04, *p* = 0.151), which also has the highest co-labeling rate of any pair in the corpus (0.21).

*Global label structure.* As an omnibus check, the silhouette coefficient of the emotion labels was computed on the 344 segments carrying exactly one label, across the seven categories with *n* ≥ 10 such segments, using cosine distance in the embedding space. The observed value was +0.0164 against a shuffled-label null of −0.0448±0.0113 (*z* = 5.43, *p <* 0.005, 200 permutations). The absolute value is low, and expectedly so: a high silhouette would indicate discrete, well-separated clusters, which is the hypothesis this paper rejects. Emotion labels are recoverable from semantic position, but they explain little of it.

*Sensitivity to the coding of structural labels.* The two structural categories are scored 0 in the valence definition (Section A.4), and they are pervasive: 655 of the 1,064 annotated segments carry at least one, and 226 carry nothing else. Because a passage such as “the insurance papers are in the drawer” is not affectively neutral in a suicide note, we tested whether the result depends on this coding (Table 1). Excluding structural labels from the valence mean, which drops the 226 structural-only segments, gives *I* = 0.1908 (*z* = 19.47), slightly *higher* than the published value. Reducing valence to its sign, discarding ties, gives *I* = 0.2161 (*z* = 19.75). Dispensing with valence altogether and testing positive-affect and negative-affect membership as separate binary concentration statistics, both are strongly concentrated (positive, *n* = 472, *z* = 12.37; negative, *n* = 650, *z* = 9.56; both *p <* 0.001). The gradient is therefore not an artifact of treating structural language as neutral; if anything, the neutral coding attenuates it by pulling mixed segments toward zero.

### **A.6** Cross-encoder robustness

We recomputed the spatial-autocorrelation statistic and the per-emotion concentration tests using each of the three encoders described in Section A.2, in each case within that encoder’s own embedding space rather than in any projection.

All three recover the valence autocorrelation at closely comparable magnitude (*I* = 0.1848, 0.1439, 0.1387), despite differing in architecture, pretraining domain, pretraining objective, and embedding dimension (Table 2). The rank ordering of per-emotion effect sizes is preserved: the same categories concentrate most strongly under each encoder (Table 3). Two categories differ appreciably between encoder families. *Information* and *happiness_peacefulness* concen-trate more strongly under the two RoBERTa models than under MiniLM, and *guilt* concentrates more strongly under MiniLM. The love–hopelessness asymmetry that motivates the Discussion is present in all three.

### **A.7** Stability of the display projection

Although no statistic depends on the projection, the figures do, so we verified that the projection is not idiosyncratic to one random initialization. We generated 20 UMAP embeddings of the same MiniLM vectors, varying only the seed, and computed pairwise orthogonal Procrustes disparity [18, 21] between each of the ^(20)^ = 190 pairs after rigid alignment, against a shuffled-rows null. Mean disparity was 0.022 ± 0.007 (range [0.011, 0.040]) against a shuffled-rows null of 0.999, indicating that UMAP recovers essentially identical structure across initializations up to rigid transformation. Recomputing Moran’s *I* on each projection gives 0.162 ± 0.009 across the 20 seeds, all significant at *p <* 0.001.

### **A.8** Sensitivity to de-identification

The corpus is de-identified by systematic substitution: personal names are replaced with a small set of substituted given names and surnames, street addresses with a single substituted address, and telephone numbers with two substituted numbers. The substitution is uniform, so the replacement tokens recur at high frequency: at least one appears in 1,001 of the 1,243 segments (80.5%), with 5,065 occurrences in total. An encoder has no way to know these tokens are artifacts, and because they are shared across most of the corpus they could plausibly organize semantic neighborhoods and thereby shape every geometric statistic reported here.

We tested this directly. Every segment was re-encoded with the substitution tokens removed, using the same encoder and settings, and all headline statistics were recomputed in the resulting space. As a control, the 242 segments containing no substitution token were unchanged under re-encoding (cosine similarity to the original embedding = 1.0000), confirming that the encoder is deterministic and that any observed displacement is signal rather than run-to-run noise. The perturbation is substantial for the segments that do contain such tokens: mean cosine similarity to the original embedding is 0.89, with a minimum of 0.50.

Every reported statistic increases when the substitution tokens are removed (Table 4). The substitution tokens therefore dilute the affective structure rather than create it, which is what one would expect of an affectively neutral signal distributed across four fifths of the corpus. We retain the original embeddings as primary, because they correspond to the corpus as it exists and as it is made available to other investigators. Removing the tokens strengthened every headline statistic, so the structure reported here comes from the language of the notes and not from the de-identification tokens, and the reported effects are, if anything, understated. Because systematic token substitution is standard practice in clinical text corpora, this bias is unlikely to be specific to this dataset, and its magnitude across corpora and embedding models is a question for subsequent research.

**Table 4:** Table S4. Headline statistics before and after removal of de-identification substitution tokens. All values computed in the original 384-dimensional MiniLM space on a cosine *k* = 15 nearest-neighbor graph. Every statistic increases when the substitution tokens are removed and the segments re-encoded.

| Statistic | Original | Tokens removed |
| --- | --- | --- |
| Valence Moran’s $I$ | 0.1848 | 0.1933 |
| Valence Moran’s $z$ | 19.68 | 20.93 |
| *Love* concentration $z$ | 15.74 | 16.81 |
| *Hopelessness* concentration $z$ | 10.81 | 12.21 |
| Emotion-label silhouette $z$ | 5.43 | 6.55 |

### **A.9** Exploratory analysis of sub-structure within hopelessness

The Discussion advances, as a hypothesis, that *hopelessness* is a clinically overloaded label absorbing several distinct constructs. We tested this exploratorily on the 412 hopelessness-labeled segments (368 notes) in the original embedding space. The analysis is exploratory in the strict sense: it was not pre-specified, and we report it because a hypothesis about internal structure should be accompanied by an attempt to find that structure, whatever the outcome. *Hopelessness does not partition into discrete sub-types.* HDBSCAN [23], run on the cosine-normalized embeddings with a minimum cluster size of 20, returned no clusters and classified all 412 segments as noise. *k*-means silhouette scores across *k* = 2 to 6 peaked at 0.091 (*k* = 3), low enough to indicate that any partition is imposed rather than found. On the evidence available here, the hopelessness region is a single continuous territory, not a set of separable sub-populations.

*It does, however, vary continuously along an interpretable gradient.* Taking the *k* = 3 partition as a descriptive device rather than a claim about discrete groups, and characterizing each sub-region by log-odds lexical enrichment with an informative Dirichlet prior [22] and by the cosine similarity between the sub-region’s centroid and the centroid of the *love*-labeled windows in the original embedding space: one sub-region (*n* = 153) is marked by terms of address, endearment, apology, and forgiveness; it lies very close to the *love* territory (mean cosine similarity 0.978) and is co-labeled *love* in 52% of its segments, against a rate of 32% across hopelessness as a whole. A second (*n* = 140) is marked by terms of illness, physical deterioration, and elapsed time; it lies substantially further from *love* (mean cosine similarity 0.820) and is co-labeled *love* in 24% of its segments. The third (*n* = 119) is dominated by the de-identification substitution tokens and by logistical vocabulary, lies closest to the structural categories (mean cosine similarity 0.966 to *information*, 0.959 to *instructions*), and is co-labeled *instructions* in 73% of its segments; we regard it as an artifact of the substitution procedure (Section A.8) combined with the practical content that accompanies it, and we draw no affective conclusion from it.

The gradient between the first two sub-regions, from hopelessness expressed in relational terms to hopelessness expressed in somatic and durational terms, is consistent with the hypothesis in the Discussion, and its relational pole carries the surface features that the Interpersonal Theory of Suicide associates with perceived burdensomeness [4]. We emphasize what this analysis does and does not support. It does not identify burdensomeness or belongingness in these passages because the corpus is not annotated for them. It does not establish discrete sub-types, and the clustering evidence argues positively against them. What it supports is the weaker claim that variation within hopelessness is continuous and lexically structured and that the structure runs along an axis a dimensional theory would predict. Confirmatory work requires the dedicated annotation study described in the Discussion.

### **A.10** Software

All analyses were conducted in Python 3.11 using sentence-transformers, umap-learn (v0.5.12), scikit-learn, hdbscan, and scipy. The analysis code is available from the corresponding author for inspection; as noted under Code availability, independent re-execution requires the restricted corpus.

## Notes

### Competing Interest Statement

The authors have declared no competing interest.

### Author Declarations

The Institutional Review Board of Cincinnati Children's Hospital Medical Center gave ethical approval for this work (protocol 2016-1505).

